# A Systematic Review of Peruvian Contributions to Scientific Publications on Experimental Research Against COVID-19

**DOI:** 10.1101/2023.05.03.23289455

**Authors:** Katiusca Coronel-Monje, Mayron Antonio Candia-Puma, Juan Jeferson Vilca-Alosilla, Luis Daniel Goyzueta-Mamani, Herbert Mishaelf Aguilar Bravo, Jorge Augusto Sánchez Zegarra, Haruna Luz Barazorda-Ccahuana, Eduardo Antonio Ferraz Coelho, Miguel Angel Chávez-Fumagalli

## Abstract

One of the countries most adversely affected by the COVID-19 outbreak was Peru. Worldwide scientific knowledge creation has significantly grown because of this pandemic. This systematic study aims to examine several facets of Peru’s experimental scientific production concerning COVID-19. Between December 2019 and June 2022, searches were made in the PubMed database for experimental scientific articles created in Peruvian institutions. The systematic review resulted in nine studies that meet the requirements. Data were extracted and analyzed on the type of biomedical research, the study’s applicability, the thematic area and specific thematic, journal impact factor and quartile, funding, grants, and institution of affiliation for the first and correspondence authors. The results revealed that Peru needs to promote policies to boost research funding and the number of researchers to produce information that will be useful for managing diseases in the future. Yet, despite the funding provided by national organizations like National Council for Science, Technology, and Technological Innovation (CONCYTEC), there were few publications and little international collaboration. The studies that have been published focus mostly on applied research in the areas of diagnostics, sanitary products, and treatment and transmission, and they have great visibility because they are indexed in Q1 journals. This thorough study revealed Peru’s inadequate reaction to COVID-19 regarding experimental scientific research. Peruvian authorities should think about supporting the required policies to boost the number of researchers and financial aid to produce information that may be utilized to manage potential new diseases in the future.

## Introduction

Coronavirus disease 2019 (COVID-19), caused by severe acute respiratory syndrome coronavirus 2 (SARS-CoV-2), was first reported in December 2019 in Wuhan, China, and has spread worldwide, becoming a pandemic with catastrophic effects [1–3]. SARS-CoV-2 severely affects humans because it is highly transmissible and rapidly mutating [4] and is reported to have a mortality rate between 0.8-19.6% with regional variation [5,6]. Various health strategies have been applied around the world, such as non-pharmacological interventions (use of masks, social distancing, monitoring of infected persons, etc.) and vaccination to reduce the spread of the virus and contagion [7]. However, since the emergence of SARS-CoV-2, there have been approximately 755 million cases of COVID-19 and 6.8 million deaths by February 2023 [8].

The first case of COVID-19 in Peru was reported on March 6, 2020, and community transmission began on March 17, 2020, [8]. At the beginning of the pandemic, the Peruvian government determined prevention measures and mandatory social isolation to control the spread of SARS-CoV-2 [9,10]. However, it could not avoid being one of the countries most affected in the number of cases, deaths per million, and total excess samples by the COVID-19 pandemic [11,12]. In the first half of 2021, the Lambda variant of SARS-CoV-2 became the most predominant in Peru’s Coastal and Andean regions, while Gamma predominated in the Amazon [13]. Until February 2023, the Ministry of Health of Peru reported 4.4 million positive cases and 219 269 deaths [14].

Faced with the current health crisis due to the COVID-19 pandemic, there has been an increase in scientific production on the subject worldwide in different fields due to the need to effectively control the disease (finding health solutions, treatments, diagnostic methods, understanding the pathophysiology of the virus, research into vaccines, etc.) [15,16]. As a result, the Peruvian government issued a supreme decree to encourage clinical trials on the prevention, diagnosis, and treatment of COVID-19 [17,18]. Additionally, the National Council for Science, Technology, and Technological Innovation (CONCYTEC), called for funding for research [19].

One Health can be used to treat a public health issue affecting people, animals, and the environment [17,18]. The COVID-19 pandemic has shown the importance of worldwide cooperation in developing and distributing vaccines and treatments and exchanging knowledge and resources. [19]. Additionally, scientists from several fields, including epidemiology, virology, animal health, and environmental health, collaborate as part of the One Health concept. [20]. Thus, various countries have carried out an internal investigation to respond to their own needs regarding COVID-19 according to their capacities and infrastructure [21–24]. Therefore, the objective of this research is to evaluate the generation capacity of experimental research carried out in Peru, which will help in making future decisions, both to establish future studies, to elucidate the lack of studies in certain areas, as well as to determine the country’s roadmap in a current and future state of emergency.

## Methods

### Study Protocol

The present systematic review was carried out as per the guidelines of the Preferred Reporting Items for Systematic Reviews and Meta-Analyses (PRISMA) [25]. The protocol for this systematic review was registered on INPLASY (INPLASY202340080) and is available in full at inplasy.com (https://inplasy.com/inplasy-2023-4-0080/). The systematic review has been elaborated according to PRISMA 2020 checklist (Table S1) [25].

### Search strategy

The search was limited to studies published from December 2019 to June 22, 2022, in the PubMed database (https://pubmed.ncbi.nlm.nih.gov/), with free electronic access that contains more than 35 million citations and abstracts of biomedical literature that includes various literature resources of the National Library of Medicine (NML) such as MEDLINE, PMC, and other databases [26]. The search for the terms associated in the literature with COVID-19 and Peru was carried out using the MeSH term, and the results were analyzed in a co-occurrence network map of MeSH terms in the VOSviewer software (version 1.6.18) [27]. MeSH terms (Medical Subject Headings), which are used to index the citations since this is a vocabulary controlled by the NML, organize their descriptors hierarchically so that more specific articles can be found from a broad search. Specialists from various areas constantly update the MESH term; every year, new concepts are modified and added [28,29]. The search string used in PubMed was: ((COVID-19[MeSH Terms]) OR (SARS-CoV-2 [MeSH Terms])) AND (PERU).

### Selection criteria and data extraction

The studies included in the systematic review were selected in three stages. First, duplicate articles, original articles other than the English language, critical and systematic reviews, meta-analyses, and publications other than an original article were excluded: letters to the editor, commentary, editorial and case reports, data studies, news, conference, and directory; all these classifications were considered using the PubMed filters [30–33]. Secondly, the titles and abstracts of the studies selected using the search strategy were analyzed. Finally, potentially relevant complete studies were retrieved and detached from the articles with a title or abstract that did not provide appropriate data to be considered within the systematic review. The included studies were those that were published in journals of quartile one or two, had first authors and/or corresponding authors with Peruvian institutional affiliations, and/or had Peruvian funding. The research topics for these studies had to produce new scientific knowledge, development, innovation, and/or adaptation of new or improved low-cost technologies, products, mechanisms, or services [33,34].

The following data were extracted: first author, first author’s institution of affiliation, first author’s country, corresponding author, corresponding author’s affiliation, corresponding author’s country, journal, year of publication, quartile, impact factor, institution funder, research topic, and type of research. The quartiles and impact factors of the journals were obtained from the SCImago Journal and Country Rank (https://www.scimagojr.com/) and/or on the main pages of each journal, respectively.

## Results

### Data sources and study selection

A thorough analysis of Peru’s experimental scientific research on COVID-19 was developed in the current paper. The study strategy’s flowchart was created and displayed (Figure 1). To do this, a search was conducted in the PubMed database using the search mentioned above string of MeSH terms, and a network map of the co-occurrence of MeSH terms was created. Through the search, 794 scientific papers between December 2019 and June 22, 2022, were found. A network map was created using 2,390 keywords, of which 212 achieved thresholds, and the minimum number of keyword occurrences was set at five. The most frequently occurring keywords were *“COVID-19”* (935 occurrences; total link strength: 7,327) and *“HUMANS”* (788 occurrences). The size of the nodes shows how frequently they occur. The co-occurrence of the nodes is shown by the curves connecting them in the same publication. The frequency of co-occurrences of two keywords increases with decreasing distance between nodes; in this case, the most frequent terms, such as *“COVID-19”*, *“HUMANS”*, *“SARS-COV-2”*, *“PANDEMICS”*, and *“PERU”*, are observed (Figure 2). Nine studies on experimental scientific research on COVID-19 were selected (Figure 1). In the identification, selection, and eligibility criteria, 592, 308, and 25 articles could be according to the three-step criteria. Data such as PMID, research types, the applicability of the study, theme, particular theme, year, and journal were taken out of the chosen studies (Table 1).

**Figure 1.**
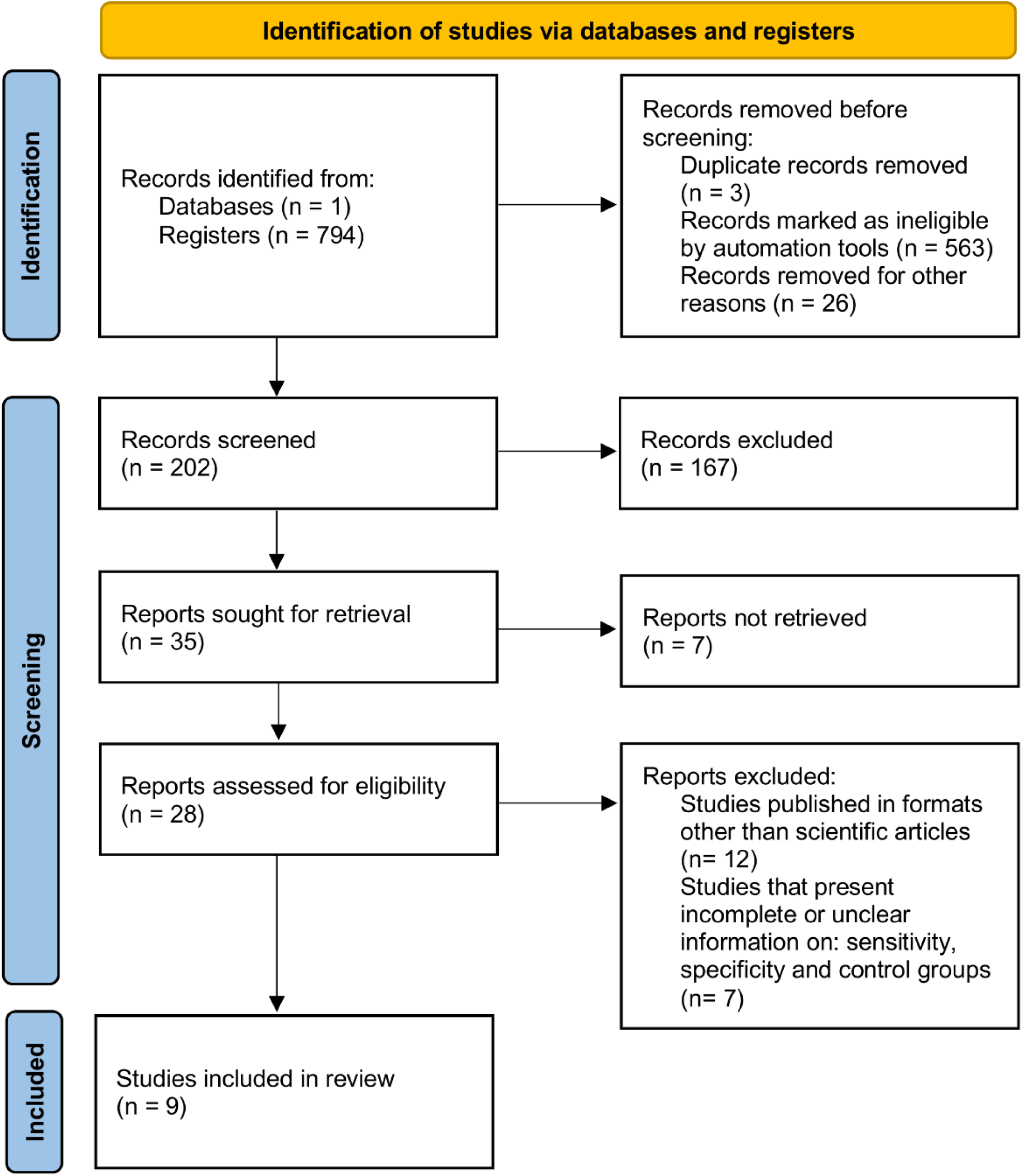
Systematic review flow diagram of the study selection process.

**Figure 2.**
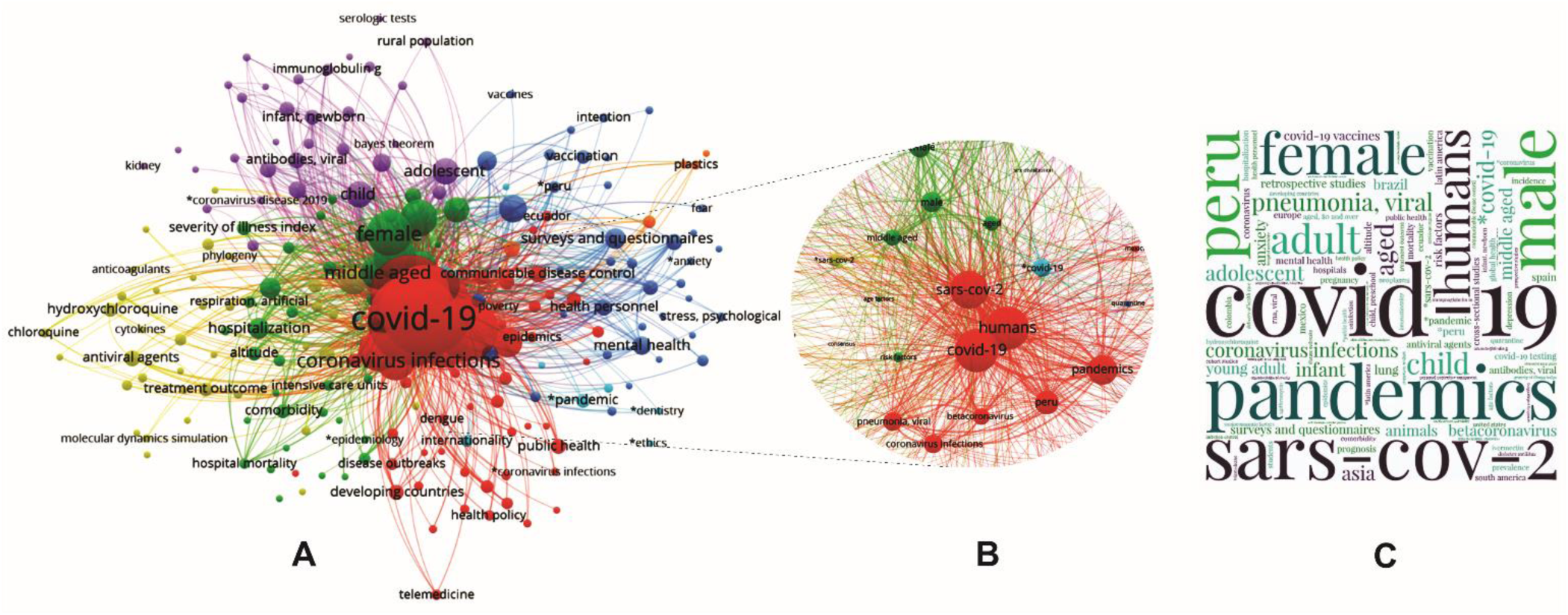
Selected articles using the PubMed database relating to COVID-19 and Peru. A) Network map built by VOSviewer based on the co-occurrence of MeSH terms. B) Red cluster zoom related to covid 19 and Peru. C) Word cloud based on the Keywords.

**Table 1.**
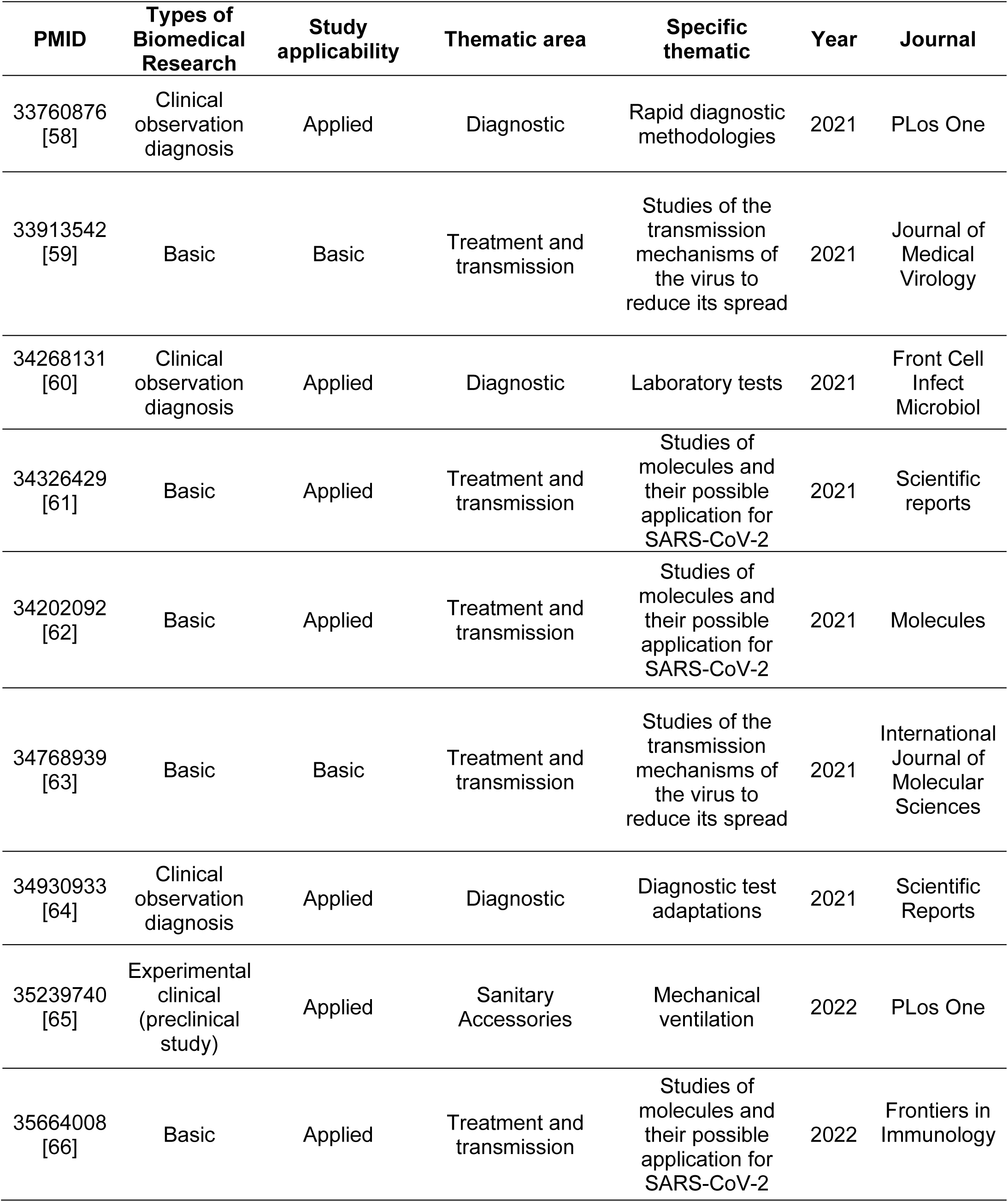
Studies on Peruvian experimental scientific research on COVID-19.

| **PMID** | **Types of Biomedical Research** | **Study applicability** | **Thematic area** | **Specific thematic** | **Year** | **Journal** |
| --- | --- | --- | --- | --- | --- | --- |
| 33760876<br>[58] | Clinical observation diagnosis | Applied | Diagnostic | Rapid diagnostic methodologies | 2021 | PLos One |
| 33913542<br>[59] | Basic | Basic | Treatment and transmission | Studies of the transmission mechanisms of the virus to reduce its spread | 2021 | Journal of Medical Virology |
| 34268131<br>[60] | Clinical observation diagnosis | Applied | Diagnostic | Laboratory tests | 2021 | Front Cell Infect Microbiol |
| 34326429<br>[61] | Basic | Applied | Treatment and transmission | Studies of molecules and their possible application for SARS-CoV-2 | 2021 | Scientific reports |
| 34202092<br>[62] | Basic | Applied | Treatment and transmission | Studies of molecules and their possible application for SARS-CoV-2 | 2021 | Molecules |
| 34768939<br>[63] | Basic | Basic | Treatment and transmission | Studies of the transmission mechanisms of the virus to reduce its spread | 2021 | International Journal of Molecular Sciences |
| 34930933<br>[64] | Clinical observation diagnosis | Applied | Diagnostic | Diagnostic test adaptations | 2021 | Scientific Reports |
| 35239740<br>[65] | Experimental clinical (preclinical study) | Applied | Sanitary Accessories | Mechanical ventilation | 2022 | PLos One |
| 35664008<br>[66] | Basic | Applied | Treatment and transmission | Studies of molecules and their possible application for SARS-CoV-2 | 2022 | Frontiers in Immunology |

### Type of biomedical research and study applicability

The type of biomedical research made the classification generally considering basic research and regarding clinical or also called applied research, specifically observational diagnostic studies, and experimental studies [34,35]. The basic biomedical research category, comprising 55.6 % (n=5) of the total studies considered in the review, came in first position as the category with the most original publications. On the other hand, clinical observation diagnosis accounted for 44.4 % (n=4) of the total number of original articles and was the following type of biomedical research with the highest number of unique publications. Of these, 11.1 % (n=1) of the total original articles of the review belonged to the type of “Experimental” clinical research, specifically a preclinical study, and 33.3 % (n=3) of the total original articles of the review belonged to the type of “Diagnostic Observational” clinical research (Table 1).

Two categories of research—basic and applied—were separated based on inquiry. Basic research seeks to understand phenomena by gathering data, whereas applied research’s major goal is to provide an accurate, practical application, or to address a particular issue. [36,37]. The systematic review results were classified according to their applicability, of which the most prevalent was “Applied” with 77.8% (n=7) of the total original articles of the review. On the other hand, the one with the lowest prevalence was “Basic” with 22.2% (n=2) of the total original articles of the systematic review (Table 1).

### Thematic area and specific thematic

The following contests, “Special Projects: Response to COVID-19” and “Special Projects: Modality Emerging Needs to COVID-19 2020-02,” both sponsored by CONCYTEC in response to the national emergency of COVID-19, proposed general and specific categories for the studies of the systematic review [33,34]. The first place was taken by the thematic area of “Treatment and transmission,” which accounted for 55.6 percent (n=5) of the total original articles. The second place went to the thematic area of “Diagnosis,” which accounted for 33.3 percent (n=3) of the total studies, while the third and final place went to the thematic area of “Sanitary accessories,” which accounted for 11.1 percent (n=1) of the total studies. Study of molecules and potential applications for SARS-CoV-2 was the specific theme that predominated the most, accounting for 33.3 percent (n=3) of all original articles, followed by “Studies of virus transmission mechanisms to reduce its spread,” accounting for 22.2 percent (n=2) of original articles. Both specific themes fall under the treatment and transmission thematic area. However, “Diagnosis” was the next most prevalent theme, with “Adaptations of Diagnostic Tests” accounting for 11.1 percent (n=1) of the total number of original articles, “Rapid Diagnosis Methodology” coming in second with 11.1 percent (n=1), and “Tests Laboratory” coming in third with 11.1 percent (n=1) of the total number of original articles (Table 1). Finally, the specific theme with the lowest number of original articles was “Sanitary accessories”, having only one specific subject and one original article “Respirators and fans” with 11.1% (n=1) of the total original articles of the review (Table 1).

### Journal impact factor and quartile

Q1 journal has a high impact factor and number of citations in a specific thematic area, allowing greater visibility of published articles [38,39]. Scientific Reports and PLoS One had the highest output rates (n=2; 22.2%), whereas only one original paper (11.1%) was published in each of the other five journals. One journal from the United Kingdom, two from the United States, and four from Switzerland made up the total number of journals in the systematic review research. Seven journals came from the first quartile (Q1). The journals with the highest production were Scientific Reports and PLoS One (n=2; 22.2%). On the other hand, the other five journals had original articles (11.1%) respectively. Four of the total journals in the systematic review studies were from Switzerland, two from the United States, and one from the United Kingdom. Seven journals were from quartile one (Q1). With the highest impact factors (20.693, 6.429, and 6.208, respectively), the Journal of Medical Virology, Frontiers in Immunology, and International Journal of Molecular Sciences are first, second, and third position, respectively. On the other side, the fourth- and fifth-placed journals are Scientific Reports with 4,996 and Molecules with 4,927. Finally, Frontiers in Cellular and Infection Microbiology and PloS One, which had impact factors of 4,300 and 3.240, respectively, were sixth and seventh in the ranking as the journals with the highest impact factor of the systematic review (Table 2).

**Table 2.** Journal impact factor and quartile of the systematic review studies.

| N° | Journal | Country | Impact Factor | Quartile | n |
| --- | --- | --- | --- | --- | --- |
| 1 | PLoS One | EEUU | 3.240 | 1 | 2 |
| 2 | Scientific Reports | United Kingdom | 4.996 | 1 | 2 |
| 3 | Frontiers in Immunology | Switzerland | 6.429 | 1 | 1 |
| 4 | Front Frontiers in Cellular and Infection Microbiology | Switzerland | 4.300 | 1 | 1 |
| 5 | Journal of Medical Virology | EEUU | 20.693 | 1 | 1 |
| 6 | International Journal of Molecular Sciences | Switzerland | 6.208 | 1 | 1 |
| 7 | Molecules | Switzerland | 4.927 | 1 | 1 |
| **Total** |  |  |  |  | **9** |

n = number of articles

### Funding and grants

Only one paper (11.1%) of the total articles was jointly funded by funds from Peru and the United States. With 22.2 percent (n=2) of the total publications in the systematic review, the National Health Institute (INS), CONCYTEC, and Universidad Católica de Santa Maria were the organizations that funded the most original articles. The country that financed the largest number of original systematic review studies was Peru, with 88.9% (n=8) of the total articles. Only one article (11.1%) of the total articles was co-financed by grants from the United States and a grant from Peru. The institutions that financed the largest number of original articles in the systematic review were the INS, CONCYTEC, and the Universidad Católica de Santa María with 22.2% (n=2) of the total articles of the systematic review, each. Co-financing for one study (11.1%) in the systematic review came from INS and CONCYTEC. On the other side, “Universidad Católica de Santa Mara” and “US Grants” jointly funded 1 study (11.1%) of the systematic review. Another study (11.1%) that CONCYTEC and Universidad San Juan Bautista jointly funded was also conducted. Only two universities in Peru—Universidad Católica de Santa Mara and Universidad San Juan Bautista—funded articles for the systematic review (Table 3).

**Table 3.** Funds and grants of the systematic review studies.

n = number of articles
| **Ranking** | **Financing Institutions** | **n** |
| --- | --- | --- |
| 1 | National Institute of Health (INS) - Perú | 2 |
| 2 | National Funding for Scientific and Technological Development (FONDECYT) | 2 |
| 3 | Universidad Católica de Santa María | 2 |
| 4 | National Institute of Health and FONDECYT | 1 |
| 5 | CONCYTEC and Universidad San Juan Bautista | 1 |
| 6 | Grant from U.S.A. and Universidad Católica de Santa María | 1 |
| **Total** |  | **9** |

### Institution of affiliation first author

Meanwhile, Universidad Católica de Santa Mara, Universidad Cayetano Heredia, and FARVET SAC are tied for second position with 22.2 percent each (n=2). The institutions with the lowest production of original papers from the systematic review are Harvard Medical School, Massachusetts General Hospital, and Pontificia Universidad Católica del Peru, all of which produced 11.11 percent (n=1) of all the systematic review’s original publications (Table 5). Regarding the institution with the highest production of original articles of the systematic review based on the affiliation of the first author. Universidad Católica de Santa María and INS occupy the first place, each with 33.3% (n=3), followed by the Universidad Peruana Cayetano Heredia, Pontificia Universidad Católica del Perú and Farmacológicos Veterinarios SAC (FARVET SAC), all of them with 11.1% (n=1). It was also evidenced that all the original articles of the systematic review produced by the “Universidad Católica de Santa María” (n=3) belonged to the thematic “Treatment and transmission”. Like this, “Diagnosis” was the theme shared by all the original papers included in the systematic review performed by INS (n=3). Universidad Peruana Cayetano Heredia and FARVET SAC, on the other hand, were both parts of the “treatment and transmission” theme. Pontificia Universidad Católica del Peru was the institution with the theme “Sanitary accessories,” to sum up. Based on the affiliation of the first author, Lima, Peru, produced the most original articles for the systematic review, accounting for 55.6 percent (n=5) of the total; Arequipa, Peru, produced 33.3 percent (n=3) of the total; and the city of Cusco, Peru produced the least original articles. The city of Chincha accounted for 11.1 percent (n=1) of all originals. In the same way, all the original articles of the systematic review produced by INS (n=3) belonged to the theme of “Diagnosis”. On the other hand, Universidad Peruana Cayetano Heredia and FARVET SAC, both belonged to the theme of “treatment and transmission”. Finally, one institution with the “Sanitary accessories” theme was Pontificia Universidad Católica del Perú. The city of Peru with the highest production of original articles of the systematic review based on the affiliation of the first author was Lima with 55.6% (n=5) of the total original articles of the systematic review, followed by the city of Arequipa with 33.3% (n=3) of the total original articles of the review. And finally, the city of Chincha with 11.1% (n=1) of the total number of original articles in the review (Table 4).

**Table 4.** Classification of the filiation, city, and theme of the first author.

n = number of articles
| **Institution** | **City and Country** | **n** | **Thematic** |
| --- | --- | --- | --- |
| Universidad Católica de Santa María | Arequipa, Perú | 3 | Treatment and transmission |
| INS | Lima, Perú | 3 | Diagnostic |
| Universidad Peruana Cayetano Heredia | Lima, Perú | 1 | Treatment and transmission |
| Pontificia Universidad Católica del Perú | Lima, Perú | 1 | Sanitary accessories |
| FARVET SAC | Chincha, Perú | 1 | Treatment and transmission |
| **Total** |  | 9 |  |

**Table 5.** Classification of the filiation and city of the corresponding author.

n = number of articles
| **Institution** | **n** | **% Studies** | **City** | **Country** |
| --- | --- | --- | --- | --- |
| INS | 2 | 22,2% | Lima | Peru |
| INS and Universidad Privada San Juan Bautista | 1 | 11,1% | Lima | Peru |
| Pontificia Universidad Católica del Perú | 1 | 11,1% | Lima | Peru |
| Universidad Peruana Cayetano Heredia and FARVET SAC | 2 | 22,2% | Lima and Chincha | Peru |
| Universidad Católica de Santa María | 2 | 22,2% | Arequipa | Peru |
| Massachusetts General Hospital and Harvard Medical School | 1 | 11,1% | Boston | United States |
| **Total** | **9** |  |  |  |

### Institution of the affiliation correspondence author

Most of the systematic review studies’ corresponding authors’ connections were with “Peru,” accounting for 88.9% (n=8) of the total. The United States came in second with 11.1 percent (n=1). Lima accounted for 44.4 percent (n=5) of the corresponding authors’ associations with Peru, followed by Arequipa and Chincha with 22.2 percent (n=2), respectively. With 33.3 percent (n=3) of the total, INS takes the top spot. Two of these articles list INS as the sole affiliation of the corresponding author, while the third includes both INS and Universidad San Juan Bautista. The second-place finishers, however, are Universidad Católica de Santa Maria, Universidad Cayetano Heredia, and FARVET SAC, each with 22.2 percent (n=2). Finally, Harvard Medical School, Massachusetts General Hospital, and Pontificia Universidad Católica del Peru are the organizations that produced the fewest original articles from the systematic review, accounting for a combined 11.11 percent (n=1) of all the systematic review’s original articles (Table 5).

## Discussion

Scientific research plays a significant role in preventing and controlling pathogens, such as SARS-CoV-2, which can cause pandemics, so it must be strengthened and increased to have a better response against future pathogens [40,41]. There has been a sharp rise in the number of scientific papers on the COVID-19 pandemic because of the numerous investigations that researchers from around the world have created. [42,43]. For this reason, this systematic review summarizes the experimental scientific research carried out in Peru against COVID-19 to identify and analyze trends and gaps in the experimental scientific field to guide the priorities and actions of researchers in future studies.

Peru is one of the countries most affected by COVID-19 [44,45], not only because of the vast death toll [45,46] but also because of the country’s economy [47]. Nonetheless, until 2021, Peru had allocated around 2.9 million dollars to 50 projects to develop scientific research related to COVID-19 [46]. However, as reported in this systematic review, the scientific production of original experimental articles has been deficient, since only 9 studies developed in Peru have been found, of which 6 were executed with government funds. This quantity, as said, is extremely modest considering that there were an average of 137 study articles every day in the months immediately after the virus’s unveiling, demonstrating how productively research groups throughout the world have been working, not only due to the high number of deaths [47,48]. Insufficient laboratory infrastructure and funding, a lack of professional security for scientists, a lack of policies to direct scientific projects, and political corruption contribute to low scientific productivity in developing nations like Peru [49–51].

Applied research made up 78% of the studies examined. In contrast to basic research, which aims to understand how nature functions without any other practical incentive, applied research focuses on using current knowledge to address a specific need [52]. Since applied research stresses the quick resolution of specific population problems, it is recommended that developing nations focus their investment efforts there [54]. Peru will continue to follow this pattern as long as the country lacks the resources to conduct basic and novel research. A developing nation is thought to benefit from focusing its investment efforts on applied research since it stresses the quick resolution of issues that impact the populace [53]. Obviously, Peru will continue along this path until it has the resources to fund basic and new research. RENACYT is the National Scientific, Technological, and Technological Innovation Registry of natural persons, Peruvian or foreign, who carry out science, technology, and innovation activities in Peru. According to the 2021 regulation, Peru has 4,702 active researchers in February 2023 [54]. Accordingly, it would have 147 researchers for every million people, of whom 31% labor in fields connected to the health sciences. This ratio is lower than that of other American nations like Chile, Colombia, and Mexico, where there are 400 researchers for every million people [58]. Peru’s weak scientific output would be directly tied to the country’s low researcher population.

Recent decades have seen a rise in the importance of scientific collaboration, which is more effective than individual work and increases the possibility of publishing in high-impact journals through collaborative research. The scientific partnership has increased and gained importance in recent decades because it is more efficient than individual work, increasing the potential for publication in high-impact journals through joint research [50,55]. Considering the affiliation of the first author and that of the reference author of the scientific studies that form part of this review, only one presented an international collaboration between the University of Peru and a US institution [56]. Given that international collaborations have a stronger beneficial impact than local or intra-university ones, Peru should boost its inclination for cooperation to increase the positive impact on research productivity. Peru should increase the propensity to collaborate to increase the positive impact on research productivity, considering that this positive effect is more significant in international collaborations than in domestic or intra-university ones [57].

## Conclusions

Peru is one of the countries that has funded the growth of experimental research related to COVID-19. Still, as this study indicates, there has been a low publication output compared to other countries in the region. Despite having a financial incentive. However, there was very little international collaboration in these papers. The fact that the researchers who wrote the publications reported it in renowned journals can be maintained. As a result, Peru should support the appropriate policies to increase the number of researchers and financial support to produce new information for the benefit of its citizens and to better prepare for pandemics like COVID-19 in the future. However, this study demonstrates that there was a weaker output of publications compared to other countries in the region. The worldwide collaboration of these papers was also quite limited. It can be saved that the researchers who created the publications reported it in highly regarded journals. Thus, Peru, as a developing nation, should encourage the necessary policies to boost the number of researchers and financial assistance to produce new information for the benefit of its people and to better prepare for pandemics like COVID-19 in the future.

## Author Contributions

Conceptualization: M.A.C.-P. and M.A.C.-F.; data curation: M.A.C.-P., J. J. V. A., K. C. M., and L. D. G. M.; formal analysis: M.A.C.-P. and M.A.C.-F.; funding acquisition: M.A.C.-P. E.A.F.C. and M.A.C.-F.; investigation: L. D. G. M., H. M. A. B., J. A. S. Z., H. L. B. C., and E.A.F.C.; methodology: M.A.C.-P. and M.A.C.-F.; writing—review and editing: L. D. G. M., H. M. A. B., J. A. S. Z., H. L. B. C., E.A.F.C., and M.A.C.-F. All authors have read and agreed to the published version of the manuscript.

## Institutional Review Board Statement

Not applicable.

## Informed Consent Statement

Not applicable.

## Data Availability Statement

Not applicable.

## Data Availability

All data produced in the present work are contained in the manuscript

https://inplasy.com/inplasy-2023-4-0080/

## Acknowledgments

Not applicable.

## Abbreviations

The following abbreviations are used in this study.

CONCYTEC: National Council for Science, Technology, and Technological Innovation
COVID-19: Coronavirus disease
FARVET SAC: Farmacológicos Veterinarios SAC
FONDECYT: National Funding for Scientific and Technological Development
INPLASY: International Platform of Registered Systematic Review and Meta-analysis Protocols
INS: National Health Institute
MeSH: Medical Subject Headings
NML: National Library of Medicine
NCBI: National Center for Biotechnology Information
PMC: PubMed Central
PMID: PubMed Identifier
PRISMA: Preferred Reporting Items for Systematic Reviews and Meta-Analyses
RENACYT: National Scientific, Technological and Technological Innovation Registry
SARS-CoV-2: Severe acute respiratory syndrome coronavirus 2

---

**Table S1.**
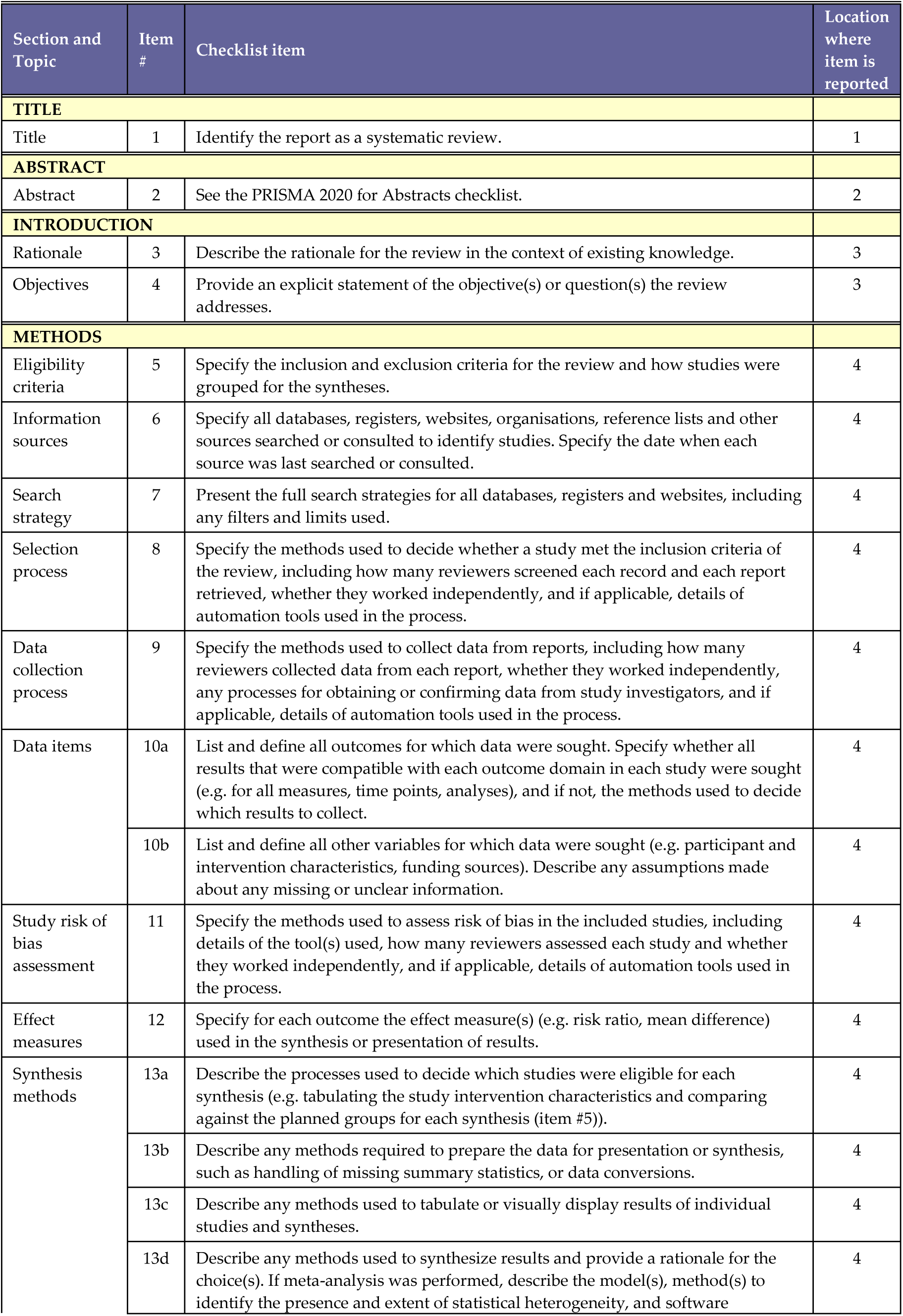
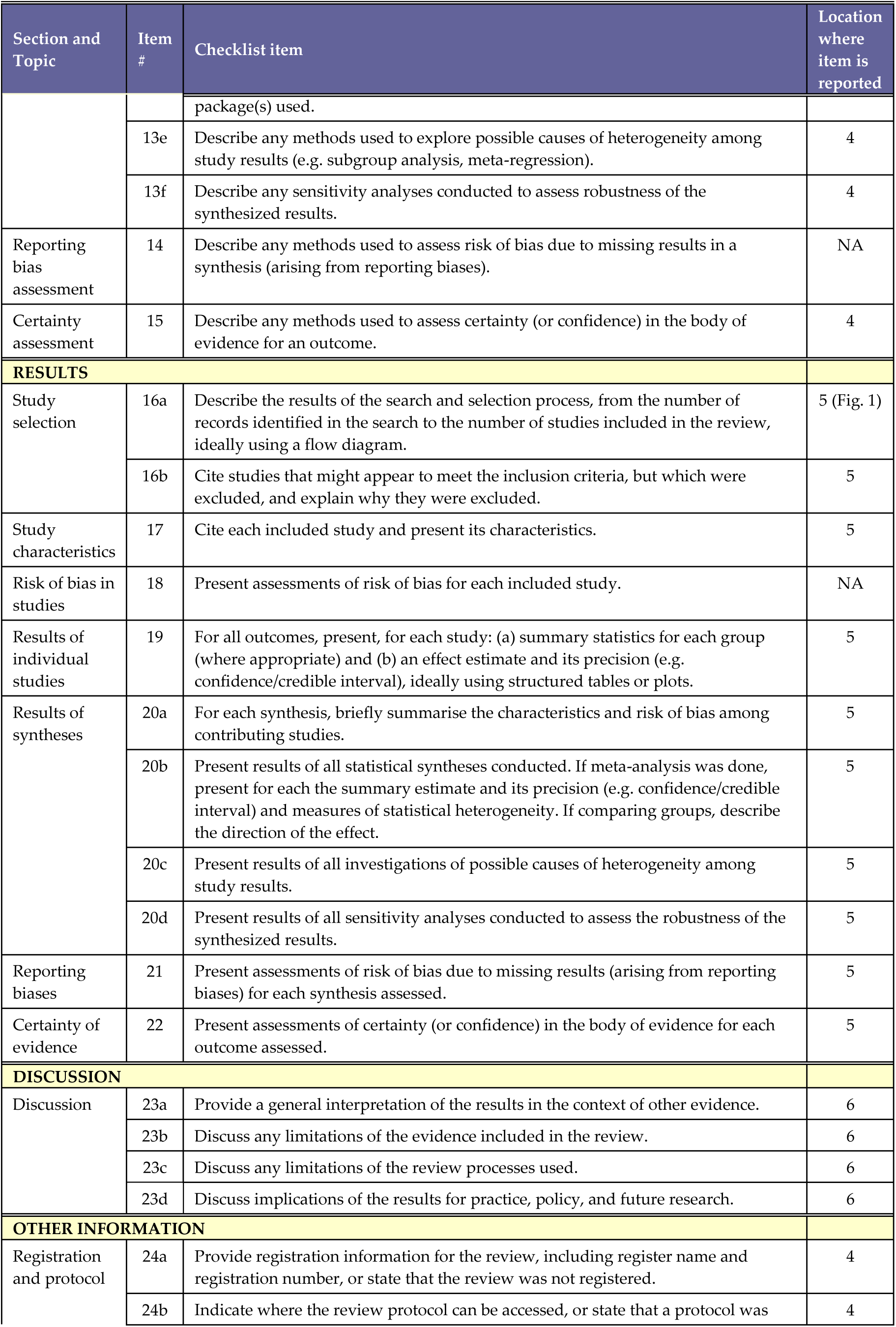
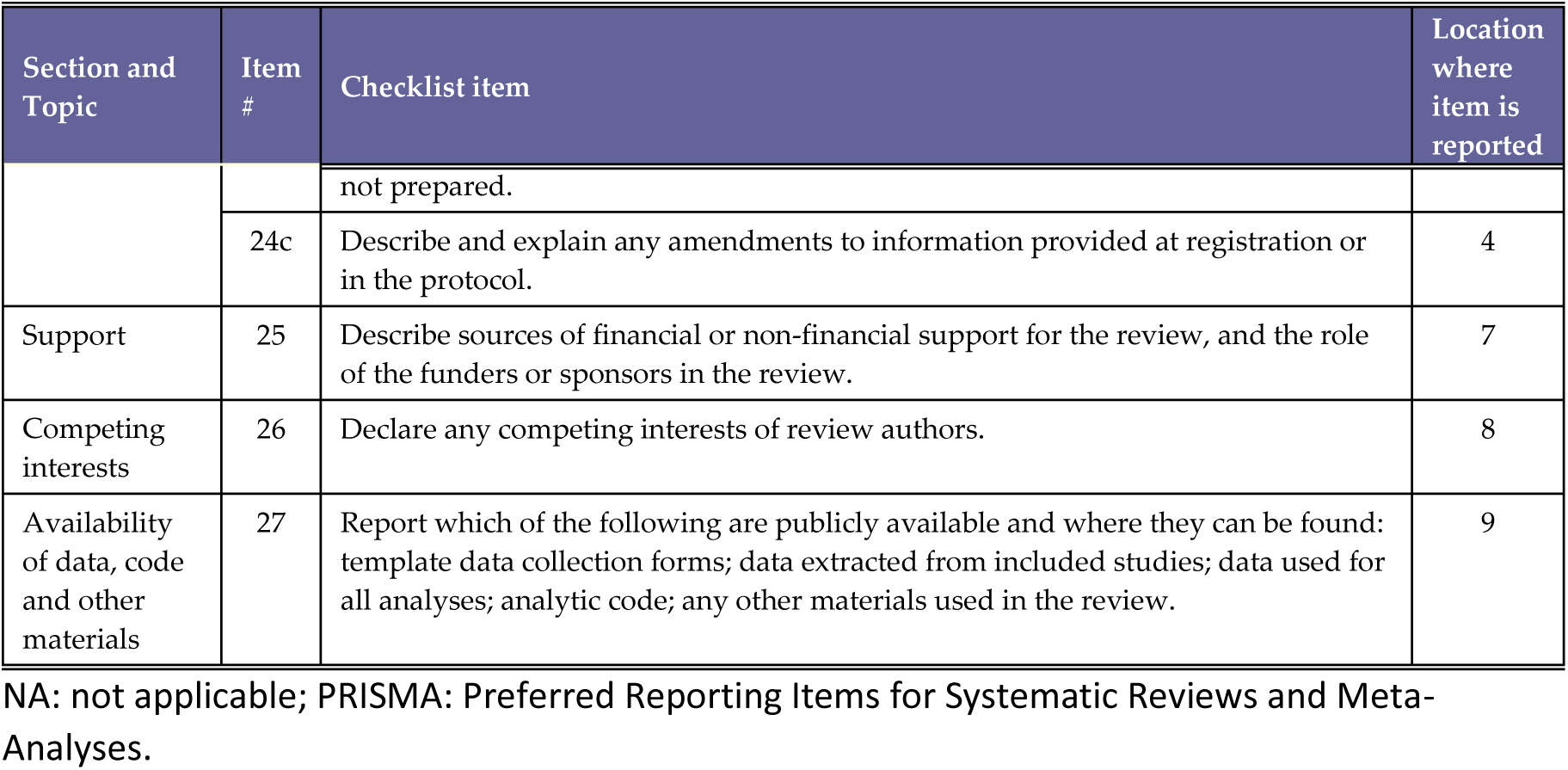
PRISMA 2020 Checklist.

n = number of articles
| Section and Topic | Item # | Checklist item | Location where item is reported |
| --- | --- | --- | --- |
| **TITLE** |  |  |  |
| Title | 1 | Identify the report as a systematic review. | 1 |
| **ABSTRACT** |  |  |  |
| Abstract | 2 | See the PRISMA 2020 for Abstracts checklist. | 2 |
| **INTRODUCTION** |  |  |  |
| Rationale | 3 | Describe the rationale for the review in the context of existing knowledge. | 3 |
| Objectives | 4 | Provide an explicit statement of the objective(s) or question(s) the review addresses. | 3 |
| **METHODS** |  |  |  |
| Eligibility criteria | 5 | Specify the inclusion and exclusion criteria for the review and how studies were grouped for the syntheses. | 4 |
| Information sources | 6 | Specify all databases, registers, websites, organisations, reference lists and other sources searched or consulted to identify studies. Specify the date when each source was last searched or consulted. | 4 |
| Search strategy | 7 | Present the full search strategies for all databases, registers and websites, including any filters and limits used. | 4 |
| Selection process | 8 | Specify the methods used to decide whether a study met the inclusion criteria of the review, including how many reviewers screened each record and each report retrieved, whether they worked independently, and if applicable, details of automation tools used in the process. | 4 |
| Data collection process | 9 | Specify the methods used to collect data from reports, including how many reviewers collected data from each report, whether they worked independently, any processes for obtaining or confirming data from study investigators, and if applicable, details of automation tools used in the process. | 4 |
| Data items | 10a | List and define all outcomes for which data were sought. Specify whether all results that were compatible with each outcome domain in each study were sought (e.g. for all measures, time points, analyses), and if not, the methods used to decide which results to collect. | 4 |
|  | 10b | List and define all other variables for which data were sought (e.g. participant and intervention characteristics, funding sources). Describe any assumptions made about any missing or unclear information. | 4 |
| Study risk of bias assessment | 11 | Specify the methods used to assess risk of bias in the included studies, including details of the tool(s) used, how many reviewers assessed each study and whether they worked independently, and if applicable, details of automation tools used in the process. | 4 |
| Effect measures | 12 | Specify for each outcome the effect measure(s) (e.g. risk ratio, mean difference) used in the synthesis or presentation of results. | 4 |
| Synthesis methods | 13a | Describe the processes used to decide which studies were eligible for each synthesis (e.g. tabulating the study intervention characteristics and comparing against the planned groups for each synthesis (item #5)). | 4 |
|  | 13b | Describe any methods required to prepare the data for presentation or synthesis, such as handling of missing summary statistics, or data conversions. | 4 |
|  | 13c | Describe any methods used to tabulate or visually display results of individual studies and syntheses. | 4 |
|  | 13d | Describe any methods used to synthesize results and provide a rationale for the choice(s). If meta-analysis was performed, describe the model(s), method(s) to identify the presence and extent of statistical heterogeneity, and software package(s) used. | 4 |
|  | 13e | Describe any methods used to explore possible causes of heterogeneity among study results (e.g. subgroup analysis, meta-regression). | 4 |
|  | 13f | Describe any sensitivity analyses conducted to assess robustness of the synthesized results. | 4 |
| Reporting bias assessment | 14 | Describe any methods used to assess risk of bias due to missing results in a synthesis (arising from reporting biases). | NA |
| Certainty assessment | 15 | Describe any methods used to assess certainty (or confidence) in the body of evidence for an outcome. | 4 |
| **RESULTS** |  |  |  |
| Study selection | 16a | Describe the results of the search and selection process, from the number of records identified in the search to the number of studies included in the review, ideally using a flow diagram. | 5 (Fig. 1) |
|  | 16b | Cite studies that might appear to meet the inclusion criteria, but which were excluded, and explain why they were excluded. | 5 |
| Study characteristics | 17 | Cite each included study and present its characteristics. | 5 |
| Risk of bias in studies | 18 | Present assessments of risk of bias for each included study. | NA |
| Results of individual studies | 19 | For all outcomes, present, for each study: (a) summary statistics for each group (where appropriate) and (b) an effect estimate and its precision (e.g. confidence/credible interval), ideally using structured tables or plots. | 5 |
| Results of syntheses | 20a | For each synthesis, briefly summarise the characteristics and risk of bias among contributing studies. | 5 |
|  | 20b | Present results of all statistical syntheses conducted. If meta-analysis was done, present for each the summary estimate and its precision (e.g. confidence/credible interval) and measures of statistical heterogeneity. If comparing groups, describe the direction of the effect. | 5 |
|  | 20c | Present results of all investigations of possible causes of heterogeneity among study results. | 5 |
|  | 20d | Present results of all sensitivity analyses conducted to assess the robustness of the synthesized results. | 5 |
| Reporting biases | 21 | Present assessments of risk of bias due to missing results (arising from reporting biases) for each synthesis assessed. | 5 |
| Certainty of evidence | 22 | Present assessments of certainty (or confidence) in the body of evidence for each outcome assessed. | 5 |
| **DISCUSSION** |  |  |  |
| Discussion | 23a | Provide a general interpretation of the results in the context of other evidence. | 6 |
|  | 23b | Discuss any limitations of the evidence included in the review. | 6 |
|  | 23c | Discuss any limitations of the review processes used. | 6 |
|  | 23d | Discuss implications of the results for practice, policy, and future research. | 6 |
| **OTHER INFORMATION** |  |  |  |
| Registration and protocol | 24a | Provide registration information for the review, including register name and registration number, or state that the review was not registered. | 4 |
|  | 24b | Indicate where the review protocol can be accessed, or state that a protocol was not prepared. | 4 |
|  | 24c | Describe and explain any amendments to information provided at registration or in the protocol. | 4 |
| Support | 25 | Describe sources of financial or non-financial support for the review, and the role of the funders or sponsors in the review. | 7 |
| Competing interests | 26 | Declare any competing interests of review authors. | 8 |
| Availability of data, code and other materials | 27 | Report which of the following are publicly available and where they can be found: template data collection forms; data extracted from included studies; data used for all analyses; analytic code; any other materials used in the review. | 9 |

NA: not applicable; PRISMA: Preferred Reporting Items for Systematic Reviews and Meta-Analyses.

